# The imprinting effect of COVID-19 vaccines: an expected selection bias in observational studies

**DOI:** 10.1101/2022.11.30.22282923

**Authors:** Susana Monge, Roberto Pastor-Barriuso, Miguel A. Hernán

**Affiliations:** National Centre of Epidemiology, Institute of Health Carlos III, Madrid, Spain; Consortium for Biomedical Research in Infectious Diseases (CIBERINFEC), Spain; Consortium for Biomedical Research in Epidemiology and Public Health (CIBERESP), Spain; CAUSALab and Departments of Epidemiology and Biostatistics, Harvard T H Chan School of Public Health, Boston, MA, USA

**Keywords:** Selection bias, collider, causal inference, SARS-CoV-2, imprinting, vaccines

## Abstract

Recent observational studies have found a higher risk of reinfection with Omicron in people who received a third booster dose. This finding has been interpreted as evidence of immune imprinting of COVID-19 vaccines. We propose an alternative explanation: the increased risk of reinfection in individuals vaccinated with a vaccine booster compared with no booster is the result of selection bias and is expected to arise even if there is no immune imprinting. To clarify this alternative explanation, we describe how previous observational analyses were an attempt to estimate the direct effect of vaccine boosters on SARS-CoV-2 reinfections, an effect that cannot be correctly estimated with observational data. We use causal diagrams (directed acyclic graphs), data simulations and analysis of real data to illustrate the mechanism and magnitude of this bias, which is the result of conditioning on a collider.

## Introduction

The SARS-CoV-2 Omicron variant and subvariants have significant antigenic changes compared with both previous variants and COVID-19 vaccines used until September 2022. There is concern that past exposure to previous variants—through infection or vaccination—can alter the immunological response to an Omicron infection in such a way that the immune response to successive Omicron infections would be impaired (1,2). This so-called immune imprinting hypothesis states that a vaccine booster in individuals who later are infected by Omicron increases the risk of a second Omicron infection. If this pernicious effect of immune imprinting truly existed, recommendations for additional vaccine doses may need to be re-evaluated.

The findings of recent observational studies in Qatar show an increased risk of Omicron re-infection in individuals vaccinated with three doses of monovalent vaccines compared with two doses (3) and no increased risk of Omicron re-infection in unvaccinated individuals (4). These findings have been interpreted as supporting the immune imprinting hypothesis, which has raised concern among authorities in charge of vaccination policies worldwide.

Here we propose an alternative explanation to the findings of the observational studies: the increased risk of reinfection in individuals vaccinated with a vaccine booster compared with no booster is the result of selection bias (due to conditioning on a collider) and is expected to arise even if there is no immune imprinting.

We start by clarifying that the causal question in immune imprinting studies is the direct effect of vaccine boosters on reinfections, and that such direct effect cannot be correctly estimated with observational data. Because a useful procedure to precisely articulate a causal question is to describe the hypothetical randomized experiment—the target trial (5)—that would answer it, we first specify the target trial for the direct effect. We then use causal diagrams and data simulations to explain that the question cannot be answered with observational data, and illustrate this by replicating the bias with real-life data.

### Specification of the target trial

Consider a target trial in which the eligibility criteria are: ≥18 years of age, having received the second dose of an mRNA vaccine at least 90 days ago and having received no third dose yet, having no previous laboratory-confirmed SARS-CoV-2 infection, and not being part of a population with special vaccination recommendations (e.g., no nursing home residents, institutionalized individuals, or health-care workers). The intervention would have three components (Figure 1). First, eligible persons would be randomly assigned to 1) immediate administration of a booster (third dose) of an mRNA COVID-19 vaccine, or 2) no further vaccine doses. Second, all participants would be forced to remain uninfected for, say, 6 months after randomization. Third, we would infect all participants with Omicron at 6 months. The outcome of interest would be a laboratory-confirmed Omicron infection at least 3 months after the first Omicron infection. (A variation would be to assign a random time of infection within, say, 6 months after randomization.)

**Figure 1.**
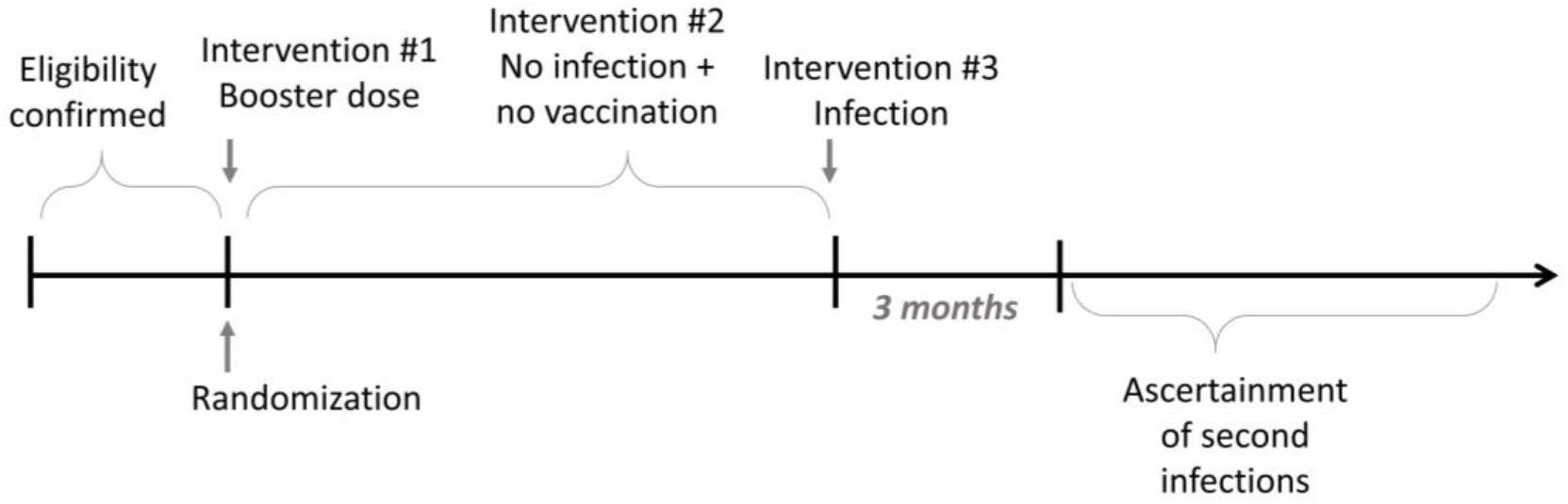
Design of a hypothetical target trial to evaluate the direct effect of a COVID-19 vaccine booster, not mediated though first infection, on the risk of reinfection.

Individuals would be followed from randomization. However, because the outcome cannot occur (by definition) until 9 months after randomization, the cumulative incidence curves for both groups would stay at zero the first 9 months. Therefore, if no reinfection can truly occur during the first 3 months after first infection and assuming there are no deaths or losses to follow-up, individuals can be equivalently followed from 9 months after randomization until the earliest of Omicron reinfection or administrative end of follow-up.

This target trial is unfeasible because, in the real world, we cannot force people to get infected at a time of our choice, but that is beside the point since the aim of this thought experiment is to specify the causal question as unambiguously as possible, rather than to design an actual trial. If this target trial could be conducted, we could use its data to quantify the (controlled) direct effect of a booster on a second Omicron infection that is not mediated through the first Omicron infection by simply comparing the risk of reinfection between individuals assigned to booster and no booster. If this direct effect exists, then there is immune imprinting.

### Emulation of the target trial is not possible

When a target trial cannot be carried out, we often use observational data from human populations to emulate it (6,7). In fact, the previous observational study (3) implicitly tried to emulate the above target trial by 1) adjusting for factors that may confound the effect of the booster on reinfection, and 2) restricting the analysis to individuals who had their first infection after the booster. Let us suppose that confounding adjustment 1) was successful and therefore the observational study appropriately accounts for the lack of randomized assignment of the booster. Even in that setting, the restriction 2) on having had a first Omicron infection is expected to introduce selection bias (8) because, in the real world, the first infection occurs more frequently among people with higher susceptibility. Therefore, if the booster prevents infections, it is essentially guaranteed that persons who received the booster and subsequently had a first infection are, on average, more susceptible to reinfection that persons who did not receive the booster and subsequently had a first infection. In the absence of data on individual susceptibility, an observational study cannot unbiasedly estimate the direct effect of a booster because restriction introduces selection bias.

To see this graphically, consider the simplified causal diagrams, which are referred to as directed acyclic graphs or DAGs, in Figure 2 (9,10). The first causal DAG represents the (randomized) target trial specified above and the second causal DAG represents the observational analysis restricted to persons with a first infection. The causal DAGs include the variables booster (yes, no), confirmed infection in period 1 (yes, no), confirmed infection in period 2 (yes, no), and an unmeasured “susceptibility” variable that represents individual characteristics that increase the risk of infection (e.g., subclinical immunosuppression, occupational and behavioral factors) or of receiving a diagnosis of infection (e.g., testing behavior, access to the health system). Period 1 ends at the earliest of 6 months or a first infection and period 2 starts right after the end of period 1.

**Figure 2.**
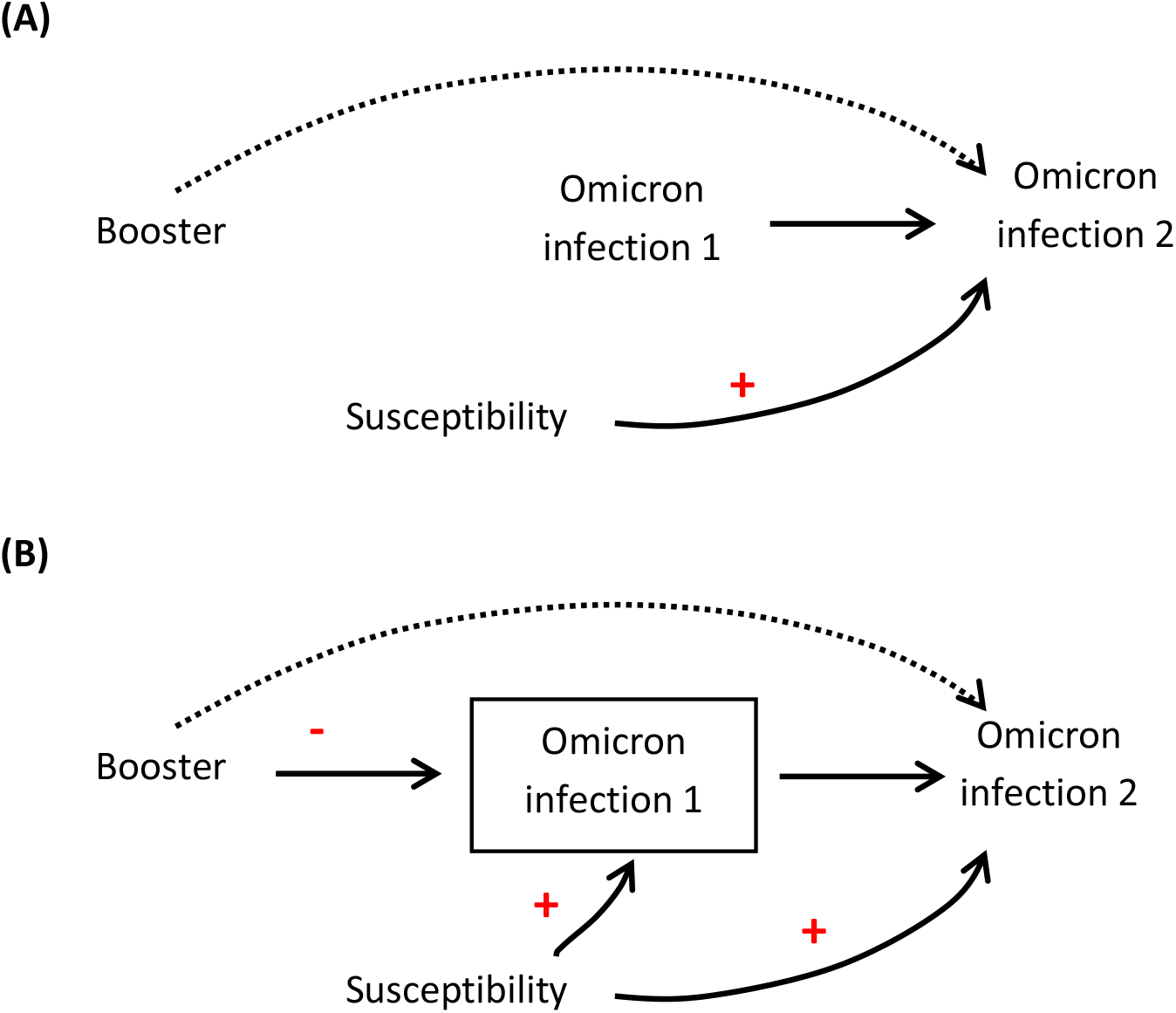
Simplified causal diagrams representing (A) a hypothetical target trial in which participants are randomly assigned to booster (yes, no) and then forced to be infected with Omicron, and (B) an observational study. The dotted line represents the direct effect of the booster, not mediated through first infection, on the risk of reinfection. The box around infection represents the conditioning of the analysis on individuals with value “yes”.

In the target trial (Figure 2A), there is no arrow from booster to infection in period 1 which, by design, occurs in all individuals, regardless of whether they were assigned to booster or no booster. In addition, there is no arrow from susceptibility to first infection because, under the intervention of the target trial, infection is guaranteed regardless of individual susceptibility. Therefore, the unconditional association between booster and infection in period 2 is an unbiased estimator of the direct effect of booster on reinfection (the dotted arrow) because everybody had a first infection. Figure 2A also represents a variation of the target trial in which individuals are randomly assigned to infection or no infection in period 1.

In the observational study (Figure 2B), there are arrows into infection in period 1 from both booster, which prevents infections (11,12), and susceptibility, which increases the risk of infection. Also, when the observational study is restricted to people with infection in period 1, we draw a box around infection in period 1 to represent the conditioning of the analysis on individuals with value “yes” on that variable. In graph theory, we say that infection in period 1 is a collider because it is a common effect of booster and susceptibility (13). Therefore, restricting to those with infection in period 1 equal to “yes” is a form of collider stratification, which is expected to induce a noncausal association between booster and susceptibility and, because susceptibility is associated with second infection, between booster and second infection (8,10,13). That is, the association between booster and reinfection among those with a first infection combines the direct effect of booster on reinfection (the dotted arrow), if any, and the selection bias induced by conditioning on first infection.

If the booster had no direct effect (i.e., if the dotted arrow did not exist) then the risk of infection in period 2 would be expected to be greater for those who did vs. did not receive the booster. This higher risk in the booster group vs. the no booster group is the entire result of selection bias and thus has no causal interpretation as a harmful effect of the booster on reinfection. In fact, all this elevated risk indicates is that people who get infected despite receiving a booster are people more susceptible to reinfection.

### Replication of the selection bias in simulated data

We designed a simplified simulation to quantify the magnitude of the selection bias under the causal DAG in Figure 2B (without an arrow from first to second infection, for simplicity). We simulated a dataset of 10 million persons with a normally distributed susceptibility variable, of whom 65% were randomly assigned to booster. We assumed that the booster decreased the probability of infection in period 1 by 50% and had no effect on the probability of infection in period 2, i.e., there was no direct effect of booster on reinfection. We considered separate scenarios in which the risk of infection is increased between 2 and 8 times per each standard deviation in susceptibility (see the Supplement for details and computer code).

For a realistic risk of infection of 10% in period 1, the odds ratio of infection in period 2 for booster vs. no booster ranged between 1.04 and 1.37, depending on the assumed distribution of susceptibility in the population. As expected, restricting the observational analysis to individuals with a first Omicron infection results in a higher risk of reinfection in the booster group even if the booster had zero effect on reinfection. It is all selection bias.

### Replication of the selection bias in real world data

We conducted the observational analysis described above using linked individual-level data from three Spanish population registries (Vaccination Registry [REGVACU], Laboratory Results Registry [SERLAB], and National Health System [NHS] registry) (12).

We identified individuals eligible for the target trial starting on January 1, 2022, when >90% of circulating variants in Spain were Omicron. We then assigned those who received a booster to the booster group and, for each of them, we randomly chose a matched control who did not receive a booster on the same week. The matching factors included sex, age (±5 years), province, time since primary vaccination (±14 days) and type of primary vaccination (BNT162b2 or mRNA-1273).

In an attempt to emulate a target trial in which all participants are infected with Omicron within 10 months of booster assignment, we restricted the analysis to individuals with a laboratory-confirmed SARS-CoV-2 infection in the next 10 months (and further matched eligible individuals on week of infection). That is, we only considered for the analysis individuals with an Omicron infection and further matched each individual in the booster group and the control on week of infection. As discussed in the previous section, restricting to individuals with infection induces uncontrollable selection bias because of differential selection between groups that depends on the unknown individual susceptibility.

We then followed individuals starting on day 90 after infection until the earliest of a confirmed SARS-CoV-2 reinfection, death, discontinuation of registration in the NHS database, or administrative censoring (October 31, 2022). To estimate the per-protocol effect, we censored at receipt of any additional vaccine dose. The cumulative incidence (risk) in each group was estimated using the Kaplan Meier method (14) and compared between groups via risk ratios (RR). Bootstrapping with 500 samples was used to compute percentile-based 95% confidence intervals (95%CI).

Of 12,749,506 initially eligible individuals, 1,704,904 experienced a first infection in the study period. Of these, 425,741 (25%) had received a booster dose before the infection. We could exactly match 249,226 (59%) individuals with a booster to the same number of controls, with a median age of 44 years. A total of 201,266 (81%) matched pairs remained under follow-up 90 days after the infection and were included in the analysis. During a maximum follow-up of 211 days (mean 133) there were 1,794 re-infections, with a 6-month risk over the full period of 0.59% in the booster group and 0.54% in the control group. The risk ratio of reinfection in the booster group compared with the no booster group was 1.08 (0.97, 1.20) at 6 months of follow-up (9 months post-infection); but varied between 1.03 (95%CI: 0.93, 1.17) in days 0 to 90 of follow-up and 1.20 (95%CI: 0.98-1.45) in days 91 to 180. As expected, the booster was associated with a higher risk of reinfection.

## Discussion

We have used causal diagrams and simulations to explain that recent observational estimates of an apparently greater risk of Omicron reinfection after a booster dose can be fully explained by the selection bias that arises when restricting the analysis to individuals with an earlier Omicron infection. We further illustrated the selection bias by conducting a real world analysis of nationwide data from Spain. Our estimates of increased risk of reinfection in individuals infected after receiving a booster was compatible with our simulation results and comparable with those from previous observational studies (3).

Removing the selection bias would require the measurement, and adjustment for, individual susceptibility to infection or diagnosis. Unfortunately, this information is not available. Comorbidities or health seeking-behavior are unlikely to fully capture individual susceptibility and, in fact, studies accounting for some of these measured factors (3) did not provide different estimates to the one in our study that accounted only for age, sex, location, and type of vaccine. Note that, even though the direct effect of booster on reinfection cannot be validly estimated and thus the presence of immune imprinting cannot be assessed with the available data, the most relevant effect to guide decision-making in vaccination programmes is the total effect of the booster on the risk of infection, which has been estimated in several observational studies (11,12,15).

Of course, other sources of bias may exist in observational studies of vaccine effectiveness, including confounding from incomplete adjustment for prognostic factors associated with vaccination, and measurement error from incomplete ascertainment of SARS-CoV-2 infection. Here we focused on the selection bias that is expected to arise in any analyses that condition on post-vaccination infection, including both analyses of observational data and of randomized trials that cannot intervene on infection itself.

In summary, analyses of observational data require a precise articulation of the causal question before the estimates can be interpreted. Observational analyses to estimate the direct effect of a booster on the risk of reinfection (i.e., “imprinting”) failed to specify the target trial that they were trying to emulate. As a result, an elevated risk of reinfection among individuals who received a booster and had a first post-booster infection was incorrectly interpreted as demonstrating a harmful effect of the booster. An explicitly causal approach to these questions indicates that 1) the elevated risk is mathematically expected and may be fully explained by selection bias and 2) observational data may not be generally used to answer these “imprinting” questions.

## Data Availability

The databases used in the observational data analysis are owned by the Ministry of Health and the Autonomous Communities in Spain, which establish the requirements for their access and use.

## Ethical statement

The use of the NHS database, REGVACU and SERLAB for the purpose of monitoring vaccine effectiveness has been approved by the research ethics committee at the Instituto de Salud Carlos III (CEI PI 98_2020 and CEI PI 08_2022). Informed consent was not required because this study is based on national population registries.

## Patient and Public Involvement statement

It was not appropriate or possible to involve patients or the public in the design, or conduct, or reporting, or dissemination plans of our research.

## Acknowledgements

We acknowledge the contribution of everyone that makes possible to have real-time data on COVID-19 vaccination and laboratory tests available in Spain, including professionals in the 19 Autonomous Communities and Cities, the Vaccines Division and Health Information Systems Department of the Ministry of Health, and the National Centre of Epidemiology at the Institute of Health Carlos III.

## Contributions

MAH and SM conceived the study and simulations, RP and SM performed the simulations, SM performed the analyses. SM is the guarantor of this article. The corresponding author attests that all listed authors meet the authorship criteria and that no others meeting the criteria have been omitted.

