## Supplementary material for "The imprinting effect of COVID-19 vaccines: an expected selection bias in observational studies"

### Supplement I. Simulation study

Susana Monge, Roberto Pastor-Barriuso, Miguel A. Hernán

#### 1. Description of the simulation

Using the causal directed acyclic graph in Figure 1B, we simulated the following variables for an observational cohort of 10,000,000 individuals:

- Susceptibility to infection $S$, a normally distributed variable with mean 0 and variance 1.

- Booster $X$, a Bernoulli variable with probability 0.65 independent of susceptibility.

- Infection in period 1 $Y_1$, a Bernoulli variable with probability given by the logistic model

  $$\log\{odds(Y_1 = 1 | S = s, X = x)\} = \log\{p/(1 - p)\} + \log(OR_s)s + \log(OR_x)x,$$

  where the expected infection risk in period 1 $p$ for average susceptibility and no booster took values 0.03 or 0.10 (short or long time periods), the conditional odds ratio for infection $OR_s$ per standard deviation increase in susceptibility took values 1, 2, 4, or 8 (no, mild, moderate, or large heterogeneity in susceptibility), and the conditional odds ratio $OR_x$ comparing individuals with and without booster was selected to numerically obtain a marginal odds ratio of 0.50 in the entire simulated population. For simplicity, we assumed no product terms.

- Infection in period 2 $Y_2$, a Bernoulli variable with probability given by the logistic model

  $$\log\{odds(Y_2 = 1 | S = s)\} = \log\{p/(1 - p)\} + \log(OR_s)s,$$

  where the expected infection risk $p$ for average susceptibility and the odds ratio $OR_s$ associated to susceptibility were assumed to be the same in periods 1 and 2. For simplicity, we assumed that neither the booster nor infection in period 1 affected infection in period 2.

#### 2. Results of the simulation

The marginal odds ratio between the booster and the infection in period 2 was null in analyses not restricted to those infected in period 1. However, as expected from the causal directed acyclic graph, conditioning on having a first infection, created a non-causal association between the booster and reinfection in period 2.

In the simulated scenarios, the upward bias in the estimated odds ratio for reinfection comparing individuals with and without booster dose increased with increasing baseline risk of infection and with increasing heterogeneity in population susceptibility:

**Table**. Estimated odds ratio for the (non-causal) association between the booster dose and a reinfection among those previously infected.

| Baseline risk of infection ($p$) | Heterogeneity in population susceptibility ($OR_s$) | | | |
|---|---|---|---|---|
| | 1 | 2 | 4 | 8 |
| 0.03 | 0.98 | 0.98 | 1.13 | 1.31 |
| 0.10 | 1.00 | 1.04 | 1.18 | 1.37 |

PRE-PRINT

### 3. R script for the simulation

```
set.seed(6482714)

# Simulated distributions for susceptibility and booster
s <- rnorm(10000000)
x <- rbinom(10000000,1,0.65)

# Simulation parameters for baseline infection risk and susceptibility
# effect in the different scenarios
p <- c(0.03,0.10)
OR.s <- c(1,2,4,8)

# Conditional odds ratios OR.x resulting in a marginal odds ratio of 0.5
# for the different combinations of p and OR.s
OR.x <- matrix(c(0.50,0.495,0.46,0.385,
                 0.50,0.485,0.43,0.35 ),nrow=2,byrow=T,
               dimnames=list(c("p.03","p.10"),
                             c("OR.s.1","OR.s.2","OR.s.4","OR.s.8")))

# Simulated infections in periods 1 and 2
mOR1 <- mOR2 <- mu.inf0 <- mu.inf1 <- OR.reinf <- adjOR.reinf <-
        matrix(NA,nrow=2,ncol=4,dimnames=dimnames(OR.x))
for(i in 1:2){
  for(j in 1:4){
    y1 <- rbinom(10000000,1,1/(1+exp(-(log(p[i]/(1-p[i]))+
                                       log(OR.s[j])*s+log(OR.x[i,j])*x))))
    y2 <- rbinom(10000000,1,1/(1+exp(-(log(p[i]/(1-p[i]))+
                                       log(OR.s[j])*s))))
    mOR1[i,j] <- sum(y1==1&x==1)/sum(y1==0&x==1)/
                 (sum(y1==1&x==0)/sum(y1==0&x==0))
    mOR2[i,j] <- sum(y2==1&x==1)/sum(y2==0&x==1)/
                 (sum(y2==1&x==0)/sum(y2==0&x==0))
    mu.inf0[i,j] <- mean(s[y1==1&x==0])
    mu.inf1[i,j] <- mean(s[y1==1&x==1])
    OR.reinf[i,j] <- sum(y1==1&y2==1&x==1)/sum(y1==1&y2==0&x==1)/
                     (sum(y1==1&y2==1&x==0)/sum(y1==1&y2==0&x==0))
    adjOR.reinf[i,j] <- exp(summary(glm(y2~x+s,
                                        family=binomial(link="logit"),
                                        subset=y1==1))$coeff[2,1])
    }
  }

# Marginal odds ratios between booster and infection in periods 1 and 2
round(mOR1,2)
round(mOR2,2)

# Mean difference in susceptibility among those infected in period 1
# with and without booster
round(mu.inf1-mu.inf0,3)

# Estimated odds ratio between booster and reinfection
round(OR.reinf,2)

# Susceptibility-adjusted odds ratio between booster and reinfection
round(adjOR.reinf,2)
```

PRE-PRINT

### Supplement II. Details of the analysis with real-world data.

**Table 1. Baseline characteristics of the total eligible population and of the matched study population for the estimation of vaccine effectiveness of the booster against laboratory-confirmed SARS-CoV-2 re-infection (restricted to the population with a first infection). Spain, January 1 to October 31, 2022, n (%)**

| | | **Total eligible population** | | **Matched sample (restricted to individuals with first infection)** | |
|---|---|---|---|---|---|
| | | **Ever received a booster (N= 7,127,328)** | **Never received a booster (N= 5,622,178)** | **Booster (N= 201,266)** | **No booster (N=201,266)** |
| Age | 18-39 | 2,213,013 (31.0) | 2,953,940 (52.5) | 59,837 (29.7) | 62,408 (31.0) |
| | 40-49 | 2,081,297 (29.2) | 1,288,042 (22.9) | 77,246 (38.4) | 75,422 (37.5) |
| | 50-59 | 2,490,710 (34.9) | 926,724 (16.4) | 56,561 (28.1) | 55,546 (27.6) |
| | 60-69 | 141,052 (2.0) | 136,648 (2.4) | 2,995 (1.5) | 3,101 (1.5) |
| | 70-79 | 136,235 (1.9) | 164,000 (2.9) | 3,523 (1.8) | 3,680 (1.8) |
| | 80-89 | 53,897 (0.76) | 109,358 (1.69) | 994 (0.49) | 998 (0.50) |
| | 90+ | 11,124 (0.16) | 43,466 (0.77) | 110 (0.05) | 111 (0.06) |
| Sex | Male | 3,654,530 (51.3) | 3,009,370 (53.5) | 108,378 (53.8) | 108,378 (53.8) |
| | Female | 3,472,798 (48.7) | 2,612,808 (46.5) | 92,888 (46.2) | 92,888 (46.2) |
| Type of vaccine for primary schedule | mRNA-1273 | 1,174,231 (16.5) | 1,246,164 (22.2) | 27,385 (16.3) | 27,385 (16.3) |
| | BNT162b2 | 5,953,097 (83.5) | 4,376,014 (77.8) | 173,881 (86.7) | 173,881 (86.7) |
| Type of booster dose | mRNA-1273 | 4,498,242 (63.1) | | 145,249 (72.2) | |
| | BNT162b2 | 2,629,086 (36.9) | | 56,017 (27.8) | |

PRE - PRINT

**Figure 1. Epidemiological week of first Omicron infection in the booster (exp=1) and no booster (exp=0) groups for the estimation of vaccine effectiveness of the booster against laboratory-confirmed SARS-CoV-2 re-infection (restricted to the population with a first infection). Spain, January 1 to October 31, 2022**

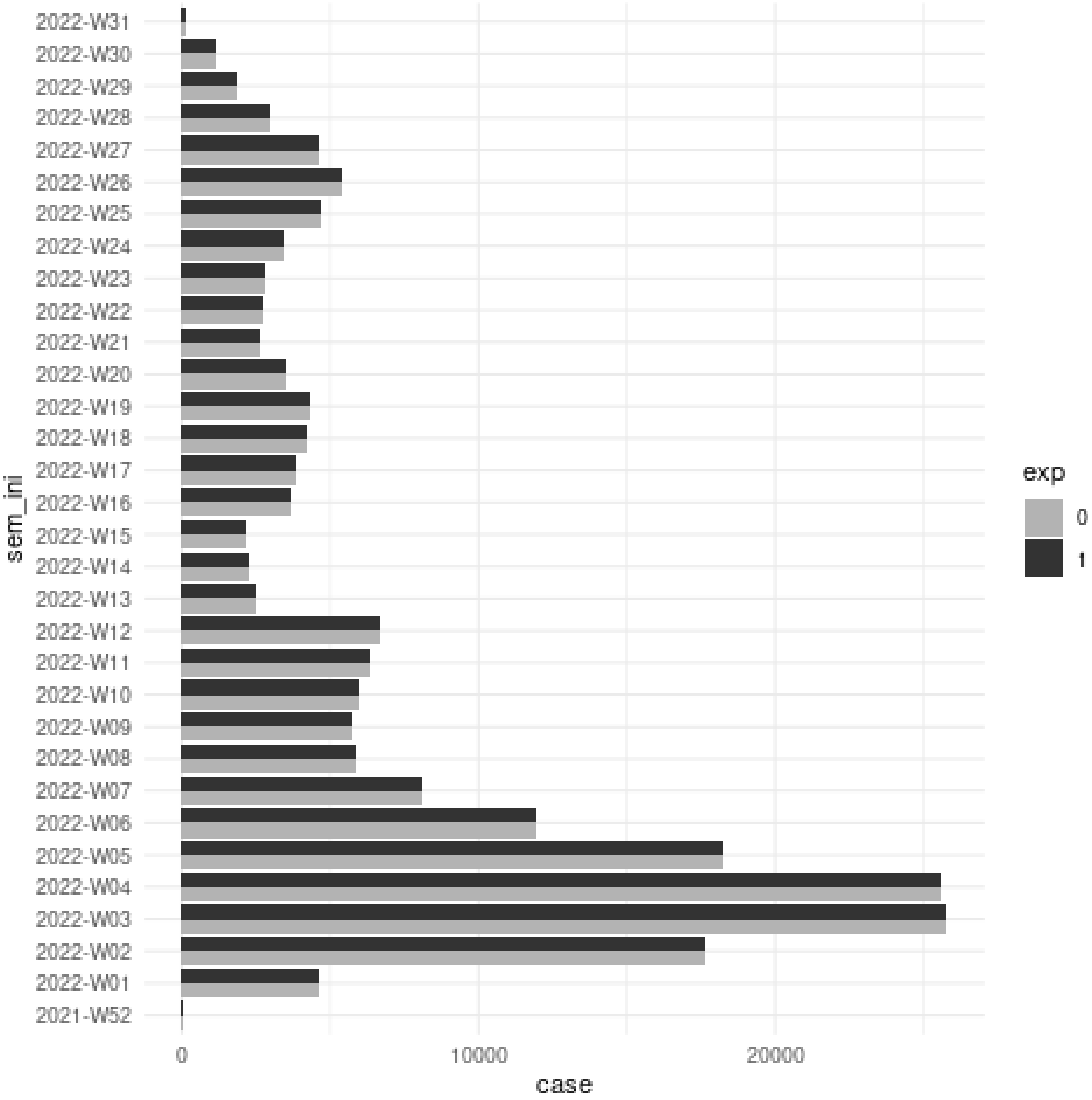
